# Breast cancer knowledge and awareness among females aged 10-24 years in Sub-Saharan Africa: A scoping review

**DOI:** 10.1101/2025.10.05.25337381

**Authors:** Punishment Peter Chibatamoto, Pisirai Ndarukwa, Moses John Chimbari

## Abstract

**Background:** Breast cancer, though rare in females aged 10–24 years, typically presents as a more aggressive disease with poorer prognosis in this population due to distinct biological features and delayed presentation. Creating awareness and disseminating knowledge of breast cancer at a young age is crucial for future risk reduction. However, levels of awareness and knowledge of breast cancer with related factors among females aged 10-24 years in Sub-Saharan Africa are not well documented. This study maps the research landscape on awareness and knowledge of breast cancer, symptoms, signs, risk factors and screening methods and identifies gaps for further research and practice.

**Methodology:** Using the Arksey and O’Malley framework for scoping studies, we reviewed literature published between 2010 and 2024. English-language articles were identified through systematic searches of PubMed, Google Scholar, and EBSCOhost. The search strategy employed a combination of keywords such as “knowledge,” “awareness,” “breast cancer,” “risk factors,” “symptoms,” “signs,” “breast self-examination,” “clinical breast examination,” “mammography,” “10-24 year old females,” and “Sub-Saharan African countries.”

**Results:** A significant research gap was identified, with only 20 studies addressing breast cancer knowledge and awareness in this specific demographic and region. The majority of the identified studies were conducted in Nigeria. While some general awareness of breast cancer exists, detailed knowledge of specific symptoms, risk factors, and breast self-examination techniques is poor across many parts of Sub-Saharan Africa. The media was frequently cited as a major source of information.

**Conclusion:** Research on breast cancer awareness, risk factors, and screening practices among females aged 10–24 in Sub-Saharan Africa is limited. The documented low levels of knowledge highlight a critical need for targeted and more effective public health interventions. Further studies are essential to investigate the underlying reasons for this knowledge gap and should be implemented across diverse settings within Sub-Saharan Africa.

**Author Summary:** We have reviewed how much is known about breast cancer awareness among young women aged 10-24 years in Sub-Saharan Africa, and we found a significant lack of research. While we know this group needs more knowledge about breast cancer and its risks to improve future outcomes, the available information is limited, with most studies focusing on Nigeria. Our review of existing literature shows that young women in Sub-Saharan Africa generally lack detailed knowledge of specific symptoms, risk factors, and breast self-examination techniques, even though media is a common source of general information. This gap highlights an urgent need for more targeted public health efforts and further studies to understand and address these issues more broadly across the region.

## Introduction

Globally, breast cancer accounted for 23.8% (2.3 million) of all cancer cases and 15.4% (670,000) of all cancer deaths in women in 2022 (Zhang et al., 2024). The developing world continues to carry a disproportionate share of breast cancer mortality of >20 per 100,000 compared to the developed world’s mortality of less than 15 per 100,00(Menon et al., 2025). The high mortality rates remain higher due to lack of knowledge and awareness of breast cancer risk factors, delayed detection, limited access to treatment, and disparities in healthcare infrastructure(Anyigba et al., 2021). In 2022, SSA accounted for 8.3% of all breast cancer cases and considerably higher mortality rates (12.5%) of the global deaths(Bray et al., 2018). Bray *et al* (2018) further observed a cumulative breast cancer incidence per 100,000 women of 38.9 in Southern Africa, 38.6 Western Africa, 30.4 in Eastern Africa and 26.8 Central Africa, and mortality rates of 15.6, 17.8, 15.4, and 15.8, respectively.

Cancer development results from a complex interplay between inherited genetic predispositions, hormonal influences, and diverse environmental factors, which include both physical and socio-economic influences. While genetics can make someone more susceptible, non-genetic factors in the physical environment, socio-economic and cultural environments significantly impact cancer risk(Shakoor et al., 2014; Calderón-Garcidueñas et al., 2005).While non-modifiable factors like age and family history exist, knowledge about how diet, physical activity, and alcohol consumption affect health allows people to make lifestyle changes to reduce risks(Łukasiewicz et al., 2021). Although developed nations have historically had higher rates of breast cancer, trends indicate rising rates in developing countries due to these lifestyle and economic changes(Porter, 2008). When women understand their individual risk factors at an early age, they can make informed decisions about their health and lifestyle to lower their risk.

Knowledge of screening methods, such as breast self-examination (BSE), clinical breast examination (CBE), and mammography is also critical. While understanding screening methods is important at any age, it is particularly critical at an early age as knowledge of these methods empowers women to proactively manage their health and facilitate timely care(Ginsburg et al., 2020). While both CBE and mammography are utilized by healthcare providers to assess breast tissue, BSE is self examination that involves both visual inspection and palpation to detect any unusual changes(Shockney, 2023). BSE complements regular mammograms and CBEs, and is not a replacement for these professional screenings. Some studies have shown that BSE is a cost-effective strategy and empowers individuals to decide on when to seek professional services(Dewi et al., 2019)(Sangwan et al., 2023).

Breast cancer is more common in women than men, with less than 1% of all breast cancer cases occurring in men due to differences in hormonal exposure, breast tissue development, and genetic predisposition(Khattab et al., 2024). Higher levels of estrogen and progesterone throughout a woman’s life are linked to an increased risk of breast cancer(Valentini et al., 2024). Although men have breast tissue, their breast ducts and lobules, where most breast cancers originate, are fewer compared to those in women. Additionally, mutations of breast cancer genes (BRCA1 and BRCA2) are more commonly associated with breast cancer in women than men. Furthermore, the risk of developing breast cancer increases with age, with 1.5% risk at age 40 years, 3% at age 50 years, and more than 4% at age 70 years(Stat bite, 2004). Over time, deoxyribonucleic acid (DNA) damage accumulates and as women age, the ability to repair this damage declines, increasing the likelihood of mutations that can lead to breast cancer(Alhmoud et al., 2020). Although over 70% of all new cases and 81% of all deaths were observed in women aged 50 years and older, the age threshold is gradually decreasing with lower survival rates amongst women diagnosed below the age of 40 years(Arnold et al., 2022).

Although invasive breast cancer is rarely diagnosed among females aged 10-24 years, it tends to be more aggressive and carries a worse prognosis in this age group, often presenting at a later stage with poorer survival outcomes compared to older premenopausal women ((Shannon & Smith, 2003). Awareness creation of breast cancer risks, signs and symptoms targeting this age group is critical for early modification of life style that may reduce the risk of developing breast cancer in the future. It has been demonstrated that 11 years is the most appropriate age at which breast topics should be introduced(Patton & Viner, 2007)(Cabrera et al., 2014). Furthermore, education on breast cancer at adolescence stage was shown to improve breast cancer outcome and survival, and promotes healthy behaviors in adulthood when breast cancer risk is greater(Brown et al., 2018)(Karayurt et al., 2008).

Thus, creating awareness of breast cancer risk factors at an early age equips young women with the ability to proactively manage their breast health, leading to better outcomes through informed lifestyle choices, regular checkups, and early detection of potential issues. It is therefore important to examine levels of breast cancer awareness and knowledge among young females under 25 years in SSA. This is essential for evidence informed strategies for the 10-24 year age group to take proactive steps towards prevention and early detection, leading to better outcomes.

We, therefore, undertook a scoping review to map the research landscape on awareness and knowledge of breast cancer, symptoms, signs, risk factors and screening methods and identify gaps for further research and practice. While scoping reviews do not employ statistical methods to compare across studies as in the case of systematic reviews and meta-analysis, they summarize the extent, range, and nature of the subject matter(Grant & Booth, 2009). This scoping review documents evidence on levels of knowledge and awareness about breast cancer, its risk factors, symptoms, signs, screening methods, sources of information and recommended interventions for improvement among females aged 10-24 years in SSA between 2010 and 2024.

## Methods

We adopted the Arksey and O’Malley’s framework for scoping studies(Arksey & O’Malley, 2005) that was enhanced by Levac, Colquhoun and O’Brien with explicit detail regarding what occurs at each stage of the review process(Levac et al., 2010) which contributed to the development of the Joanna Briggs Institute (JBI) manual(Peters et al., 2015). The JBI approach to scoping reviews comprises six stages: (1) identifying the research question, (2) identifying the relevant studies, (3) study selection, (4) charting the data, (5) collating, summarizing, and reporting the results, and the optional sixth stage of consultation with stakeholders. The sixth stage, which is optional, is consultation with stakeholderswe did not do in this review. In this paper we adhered to the PRISMA-ScR checklist in presenting the abstract, rationale, methodology, findings, and conclusions of the scoping review(Tricco et al., 2018).

In this scoping review, awareness and knowledge are defined according to the WHO Guide to Developing Knowledge, Attitude, and Practice Surveys(World Health Organization, 2008). Awareness of breast cancer involves being *generally informed* about various factors that can increase a person’s risk of developing breast cancer and knowledge encompasses a *deeper understanding* of specific risk factors and their potential impact on breast cancer development.

The following stages were followed in this scoping review:

### a) Identifying research questions

The identification of research questions provided a roadmap for data collection, analysis and interpretation stages with a focus on the relevant aspects of the research problem consistent with scoping review methodology(Peters et al., 2015). The scoping review study explored the following research questions:

- What is the nature and extent of research on knowledge and awareness about breast cancer, its signs and symptoms among females aged 10-24 years in SSA?
- What documented evidence exists on knowledge and awareness about breast cancer risk factors among females aged 10-24 years in SSA?
- What documented evidence exists on knowledge and awareness about breast cancer screening methods among females aged 10-24 years in SSA?
- What are sources of information that are most frequently studied in relation breast health among females aged 10-24 years in SSA?
- What interventions are recommended for improved knowledge and awareness of breast cancer among females aged 10-24 years in SSA?

### b) Identification of relevant studies/search strategy

We employed a three-step search strategy proposed by Joanna Briggs Institute (Peters M, Godfrey C, McInerney P, Soares CB, Khalil H, Parker D). Step one focused on limited search for already existing published research articles on knowledge and awareness of breast cancer knowledge among females aged 10-24 years in SSA, culminating in a collation of a list of relevant keywords. Step two involved a comprehensive formal search of keywords in three databases (PubMed, Google Scholar and MEDLINE hosted by EBSCO*host)* using the Boolean operators. In the third step, we used snowballing searching technique involving manually tracing the reference list of identified relevant studies for additional studies until no new information was retrieved (saturation). The keywords searched were “knowledge”, “awareness”, “breast cancer”, “symptoms”, “signs”, “breast self examination”, “clinical breast examination”, “mammography”, “risk factors”, “females aged 10-24 years”, “2010-2024” and “48 SSA countries South of the Sahara desert”. Additionally, synonyms and terms with similar definition to the above-mentioned searched terms were also employed.

### c) Inclusion/exclusion criteria for retrieving articles

Research articles included in this scoping review were selected based on the following criteria: (a) primary studies conducted in SSA (48 countries in the continent of Africa, south of the Sahara desert); (b) published between January 2010 to December 2024; (c) studies reporting evidence on famales aged 10-24 years; (d) reporting evidence on knowledge and awareness of breast cancer, symptoms, signs,screening methods and associated risk factors and primary sources of breast health related information among females aged 10-24 years in SSA and (e) articles published in English language.

The exclusion criteria were based on: (a) studies conducted in other countries but not in 48 countries of SSA, (b) studies reporting evidence in males (c) studies published before 2010 and after 2024 (d) studies published in the form of reports, editorials, conference paper, book chapters, studies involving females outside the 10-24 year agroup, and (e) studies involving non-breast cancers. Two authors independently screened the initial titles and abstracts of the retrieved articles for relevance and inclusion, and the articles that did not meet the inclusion criteria were excluded. The full-text evaluation was conducted among the remaining articles for inclusion.

### d) Charting the data

Data was extracted using a standard extraction form adapted from Udoh et al., (2020). Information on authorship, year of publication, title, study aim, country, study population, study design, key results/outcome, conclusion/recommendation was recorded on the charting form. Two authors extracted data from all the papers and cross-checked the data to reduce any possible individual bias. Authors independently extracted data, and discrepancies were resolved through brainstorming session with the third Author.

### e) Collating, summarizing, and reporting the results

This final stage of the Arksey and O’Malley methodological framework involved collating, summarizing, and reporting evidence(Arksey & O’Malley, 2005). We collated, analysed thematic data from the charting form and reported the evidence using the Braun and Clarke framework(Tricco et al., 2018). The research used a narrative approach to present each theme, ensuring that the key aspects of knowledge and awareness related to breast cancer risk factors among females aged 10-24 years were clearly communicated to the audience.

## Results

### Description of studies

Our search identified 1920 records including 77 duplicates based on PRISMA guidelines(Page et al., 2021). Subsequent to the removal of duplicates, 1772 articles were excluded as neither title nor abstract was relevant for the research questions. Following the title and abstract screeing, 71 articles were assessed for full-text eligibility, and 51 articles were excluded for not meeting inclusion criteria. Thus, 20 reports were included in the final scoping review. Figure 1 shows a the selection process of articles meeting inclusion criteria for this review.

**Figure 1:**
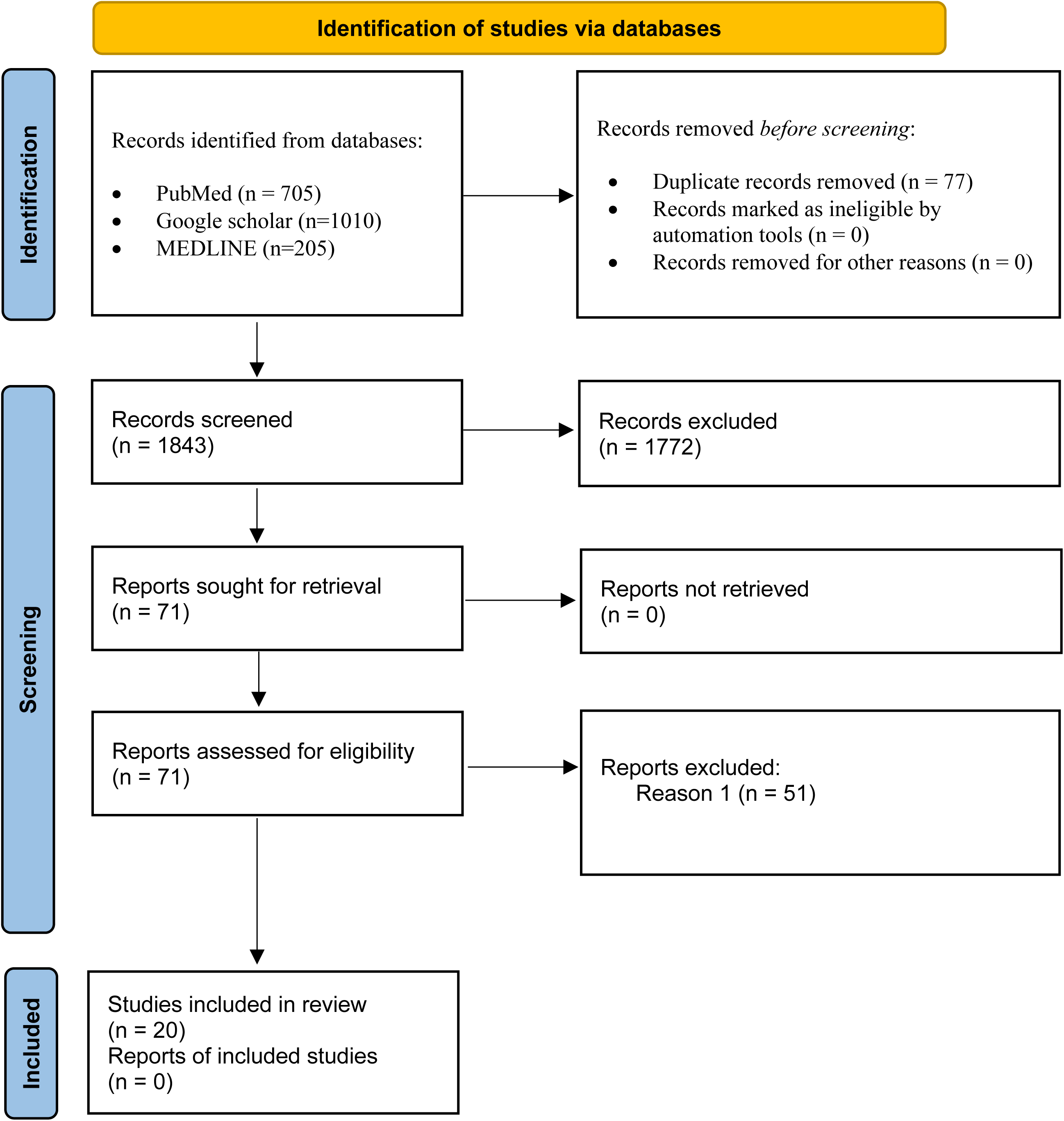
Flowchart of selection process of articles meeting inclusion criteria for this review.

### Characteristics of reviewed articles

Table 1 presents the characteristics of the 20 articles included in the scoping review process. These include identification (ID) numbers of the included articles, year of publication, authors, title of study, study aim, country of study, study population, study design, key results and recommendations. Almost half of the publications were from Nigeria (n=9: IDs 2; 9; 10; 12; 13; 15; 18; 19 and 20). Other articles were of studies conducted in Ethiopia (n=3: IDs 3; 8 and 17), Angola (n=1: ID=1), Uganda (n = 1: ID 5), Rwanda (n=1: ID 5), Cameroon (n=1: ID 7), Togo (n=1: ID11), Democratic Republic of Congo (n=1: ID 14) and Ghana (n=1: ID 16).

**Table 1:** Characteristicsof retrieved articles from Sub-Saharan African countries.

| ID # | Year | Author & Year | Title | Study Aim | Country | Study Population | Study Design | Key Results/Outcome | Conclusion/Recommendation |
| --- | --- | --- | --- | --- | --- | --- | --- | --- | --- |
| 1 | 2012 | Martha Nyanungo Sambanje, Benford Mafuvadze | Breast cancer knowledge and awareness among university students in Angola | Assess breast cancer knowledge and awareness among university students in Angola | Angola | 595 (350 females and 245 males) university students | Cross-sectional | Insufficient knowledge of breast cancer and its risk factors associated with breast cancer among university students in Angola irrespective of whether they were in medical or non-medical programs. Majority of the participants were not aware of some of the early signs of breast cancer such as change in color or shape of the nipple, even though they appreciated the need for monthly breast self-examination. Overall most of the participants indicated the need for increased breast cancer awareness among university students. | The study recommended designing health education programs on raising awareness, promoting screening and early detection of breast cancer. |
| 2 | 2013 | Iheanacho P, Ndu A and Emenike AD | Awareness of breast cancer risk factors and practice of BSE among female undergraduates in university of Nigeria Enugu campus | Determine the awareness of breast cancer risk factors and practice of breast self examination among female students of the University of Nigeria Enugu Campus | Nigeria | 240 female students at a Nigerian university | Descriptive | Most of the students have little knowledge of breast cancer risk factors and majority of the students do not practice BSE monthly | Low level of knowledge on breast cancer risk factors and BSE and only few students practice BSE monthly. There is need for more public enlightenment on breast cancer risk factors and importance of BSE especially among students. There should also be regular organization of seminars and workshops for students to address sensitive topics like breast cancer risk factors and BSE. |
| 3 | 2014 | Ameier K, Abdulie SM, Pal SK, Arebo K and Kassa GG | Breast cancer awareness and practice of BSE among female medical students in Haramaya University, Harar, Ethiopia | Assess the knowledge, attitude and practice of BSE among female undergraduate medical students of Haramaya University, Ethiopia. | Ethiopia | 126 female medical students | Cross sectional | All participants knew about breast cancer, and more than 80 % of the participants had very good knowledge of all the risk factors. However, the actual practice of BSE was very low (23%) among the medical students. | Knowledgeable of risk factors of breast cancer and BSE was high among medical students in Ethiopia. However, the practice for BSE was low. The study recommended education of practicing BSE. |
| 4 | 2014 | Peltzer K and Pengpid S | Awareness of breast cancer risk among female university students from 24 low, middle income and emerging economy countries. | Investigate the awareness of breast cancer risk factors among female university students in 24 low, middle income and emerging | Cameroon, Ivory Coast, Madagascar, Mauritius, Namibia & Nigeria | 2,126 female university students (359 Cameroon; 368 Ivory Coast; 383 Madagascar; 331 Mauritania; | Cross-sectional | 35.4% of the female students were not aware that any of these risk factors could influence breast cancer. Less than 30% of students in Ivory Coast, Madagascar and Nigeria aware of | The study revealed a low breast cancer risk perception across 8 different risk factors. Awareness raising of risk factors and breast cancer was recommended. |
|  |  |  |  | economy countries. |  | 335 Namibia and 350 Nigeria) |  | genetic causes of BC. |  |
| 5 | 2016 | Godfrey K, Agatha T, Nankumbi J. Breast | Breast cancer knowledge and BSE Practices among female University students in Kampala, Uganda: A descriptive study | Assess female university students' knowledge of breast cancer risk factors, signs and symptoms, and identify BSE practices | Uganda | 204 female university students | Cross-sectional descriptive | High awareness of breast cancer (98.0%) and BSE practices (76.5%) among female students. Over half the students (61.3%) had an intermediate level of knowledge about risk factors related to breast cancer and the signs and symptoms of the disease. Skills related to BSE practices were found to be low (43.6%). Majority (56.9%) of students received information about breast cancer via mass Media | Pre-post education intervention studies need to be conducted to evaluate the intervention outcomes related to breast cancer knowledge and BSE practices among female students in Uganda |
| 6 | 2016 | Ndikubwimana J, Nyandwi JB, Mukanyangezi MF and Kadima JN | Breast cancer and BSE: Awareness and practice among secondary school girls in Nyanugenge district, Rwanda | Assess the level of early sensitization and education of adolescent high school girls in Rwanda about breast cancer | Rwanda | 239 secondary school girls | Cross-sectional | Almost all (94.6%) of the surveyed girls had heard about BC, but less than 25% had sufficient knowledge about BC diagnostic methods, BC risk | Knowledge of BC and BSE among adolescent girls practice is low. Media plays a critical role in disseminating information on BC and BSE. However, a coordinated cooperation between public health educators, parents and the media in dissemination of |
|  |  |  |  | and BSE as one of strategic approaches to reduce the risk of late intervention and thence the BC related deaths |  |  |  | factors and BSE. More than 74% of interviewed girls never had BSE for breast cancer purpose, and of those who agreed doing BSE, only 4% knew the real timeframe of doing it, consistent with the studies by other researchers worldwide. | adequate information was lacking.. |
| 7 | 2017 | Sama CB, Dzekem B, Kehbila J, Ekabe CJ, Vofo B, Abua NL, Dingana TN, Angwafo F | Awareness of breast cancer and BSE among female undergraduate students in a higher teacher training college in Cameroon | Assess the awareness, knowledge and perceptions of breast cancer and practice of breast self-examination among female undergraduate students in a higher institution of teaching in Cameroon | Cameroon | 345 female university students. | Cross-sectional | The study indicates that there was considerable awareness about the existence of breast cancer, but insufficient knowledge and misperceptions on its risk factors and causes as well as infrequent practice of BSE | Knowledge on risk factors and clinical presentation of breast cancer was insufficient among female undergraduate students in Cameroon. The practice of BSE was also low. Educational programmes are recommended for raising knowledge levels on risk factors of breast cancer. |
| 8 | 2018 | Gebresillassie BM, | Evaluation of knowledge, perception, and risk awareness | Assess the knowledge, perception and risk | Ethiopia | 300 female medical students | Cross-sectional | Most (61.7%) scored below 50 out of 100 on overall knowledge of | Study recommended provision of comprehensive breast cancer and health awareness programs for female |
|  |  | Gebreyohannes EA, Belachew SA and Emiru YK | about breast cancer and Its treatment outcome among University of Gondar students, Northwest Ethiopia | awareness about breast cancer among female medical and health science students of University of Gondar, Ethiopia |  |  |  | breast cancer, 41% on risk factors and 66.7% on symptoms. Majority of the participants were unaware for complex risk factors such as first child after the age of 30 years, early onset of menses, and menopause after the age of 55 years. More than two third 188 (62.7%) of the participants had no previous participation in breast care awareness | youngsters. |
| 9 | 2018 | Ifediora CO and Azuike EC | Tackling breast cancer in developing countries: insights from the knowledge, attitudes and practices on breast cancer and its prevention among Nigerian teenagers in secondary schools. | Ascertain the KAP of breast cancers and BSE among female final year students enrolled at secondary schools in Anambra State, Nigeria | Nigeria | 432 female secondary students | Cross sectional | Knowledge on specific risk factors and symptoms was low (41.5% and 46.1% respectively) among female senior secondary school students. However, general knowledge of breast cancer was high (75.2%) | Study recommended incorporation of education on breast cancer into secondary school academic curricula of resource-limited countries. |
| 10 | 2019 | Ifediora CO and Azuike EC | Sustainable and cost-effective teenage breast awareness campaigns: Insights from a Nigerian high school intervention study. | Evaluate ways to ensure sustainability of an intervention of early enlightenment to empower teenage high school girls | Nigeria | 432 high school female students | Cohort study | Significant improvements on specific knowledge regarding risk factors and early symptoms of breast cancer, as well as BSE techniques and regular monthly practice. video-assisted, face-to-face and handouts were both effective and of similar impact on improvements on most aspects of knowledge regarding the breast cancer's early symptoms and risk factors after introduction of a 6 months longitudinal intervention. | The study recommended adjustments of existing high school curricula to allow repeated yearly teachings on breast cancer, along with in-built-in evaluation systems like examinations and quizzes, are ways of ensuring engagement. |
| 11 | 2020 | Darre, T., Tchaou, M., Djiwa, T., Tcharié, E.L., Brun, L.V.C., Gbeasor-Komlanvi, F.A., N'Timon, B., Amadou, A., Simgbani, P., Ekouévi, D.K. and Napo-Koura, G | Breast cancer: Knowledge, attitudes on risk factors and means of screening by medical students from Lomé, Togo | Assess knowledge and attitudes about breast cancer screening among students in the Faculty of Health Sciences at the University of Lomé. | Togo | Medical students (661 male and 273 female) | Descriptive | The students' response to the risk factors for breast cancer was generally satisfactory. | Need for further strengthening in order to better understand its risk factors |
| 12 | 2020 | Elemile MG, Ayamolowo LB, Owolabi AG, and Ighrakpata O | Awareness and practice of BSE for early detection of breast cancer among female adolescents in selected secondary schools in Ado-Ekiti, Nigeria | Assess the level of awareness and practice of BSE among senior secondary school girls in Ado-Ekiti. | Nigeria | 240 female students senior secondary | Cross sectional | 29.6% of the respondent had good level of awareness of cancer of the breast, while 73.9% did not have knowledge that breast inspection could help in the timely recognition of breast malignance. Also 49% reported that they are not conscious of how breast BSE is being carried out, 32% reported that BSE is not necessary to be done by female. The lack of awareness about the ailment could be attributed to low level of knowledge including related risk factors. | There is a need for sensitization on breast cancer information among adolescents |
| 13 | 2020 | Grace Enomfon Akpan GE and Abatai E | Knowledge of breast cancer symptoms, risk factors, preventive measures and management among students in University of Uyo | Assess the level of knowledge of breast cancer among students of the University of Uyo | Nigeria | 285 university students (142 males 143 females) | Descriptive study | Students possessed low level of knowledge of breast cancer risk factors (37.6%) and their knowledge of breast cancer symptoms and preventive measures was average (50.2% and 66.2%) respectively. Overall, knowledge of breast cancer was average. | Health education on breast cancer should be incorporated in general studies courses (GSTs) |
| 14 | 2021 | Mpulumba B, Mukendi K, Musasa K, Manongo B, Bambi N, et al. | Knowledge on breast cancer and practice of BSE among schoolgirls in Democratic Republic of the Congo | Assess knowledge about breast cancer and the practice of BSE among schoolgirls, and to propose effective intervention measures to | Democratic Republic of the Congo | 962 high school girls | Descriptive cross-sectional | School girls (61.75%) knew about breast cancer. The risk factors of breast cancer were unknown (59.56%) and for 47.56% of school girls, BSE was a preventive way, whereas 61.64% | Sensitization should be focused on knowledge, practice of breasts self-examination of an organization of screening campaigns. |
|  |  |  |  | promote early diagnosis and treatment of breast cancer in Mbujimayi |  |  |  | did not know about BSE. |  |
| 15 | 2021 | Sadoh AE, Osime C, Nwaneri DU, Ogboghodo BC, Eregie CO, Oviawe O | Improving knowledge about breast cancer and BSE in female Nigerian adolescents using peer education: a pre-post interventional study | Determine the effect of peer education as a strategy to create awareness on BC and BSE among in-school female adolescents in Benin City. | Nigeria | 1337 pre- and 1201 post female students | Pre-post intervention | Breast cancer knowledge score ($20.61 \pm 13.4$) prior to training was low and it statistically significantly improved to $55.93 \pm 10.86$ following training ($p < 0.0001$) | Peer education strategy improved knowledge on BC and BSE among adolescents. Peer education strategy is low cost and can be useful in low resource settings. |
| 16 | 2021 | Osei-Afriyie S, Addae AK, Oppong S, Amu H, Ampofo E, Osei E | Breast cancer awareness, risk factors and screening practices among future health professionals in Ghana: A cross-sectional study | Explore the awareness, risk factors, and self-reported screening practices of breast cancer among female undergraduate students at the University of Health and Allied Sciences | Ghana | 385 female university students | Cross sectional | There was moderate awareness of the modalities of breast cancer screening and the risk factors of breast cancer among the students with 73% of the students aware of breast cancer, with social media being the most important source of information (64.4%). Prevalence of breast cancer risk factors varied from 1% of having a personal history of breast cancer to 14.3% for positive family history of breast cancer. | Awareness campaigns and education should be intensified in the University to bridge this gap. |
| 17 | 2022 | Mehiret G, Molla A, Tesfaw A. | Knowledge on risk factors and practice of early detection methods of breast cancer among graduating students of Debre Tabor University, Northcentral Ethiopia | Assess the knowledge on risk factors and practice of early detection methods of breast cancer among female graduating students of Debre Tabor University. | Ethiopia | 270 female students | Cross-sectional | All of the students heard about breast cancer. about 208 (77.1%) of the respondents respond family history is a risk factor for breast cancer. | Focus on giving health information regarding risk factors of breast cancer and methods of early detection of breast cancer to the public & their clients |
| 18 | 2022 | Saulawa HT, Auwal FI and Danjuma NM | Knowledge and awareness of breast cancer risk factors and screening methods amongst undergraduate pharmacy students of Ahmadu Bello University Zaria, Nigeria | Assess knowledge and awareness of breast cancer risk factors and screening methods amongst undergraduate pharmacy students of Ahmadu Bello University (ABU) Zaria, Nigeria. | Nigeria | 89 female and 139 male university students | Cross sectional | The study revealed a fair level of knowledge and awareness of breast cancer risk factors and screening methods amongst undergraduate pharmacy students of ABU Zaria. | There is need to update the curricular of clinical pharmacy training to include a course on cancer so that pharmacists will be better informed about risk factors |
| 19 | 2023 | Magdalene E. Effiong, Israel S. Afolabi and Shalom N. Chinedu | Influence of age and education on breast cancer awareness and knowledge among women in south western Nigeria | Assess the level of awareness and knowledge of BC and BSE amongst female students and staff of six educational institutions in Ota, Southwest Nigeria | Nigeria | 917 female students | Cross sectional | Results were diasgregated into awareness and knowledge with more participants aware of BC (94.80%) than BSE (65.11%). Major sources of information were television (65.37%), health workers (45.90%) and internet (45.11%). Television and radio were most effective (62.11%), followed by social media (50.22%) and testimonies of BC survivors (46.43%). Seven participants (0.62%) have BC while 55 (4.85%) have a family history of BC although not clinically established. On the overall, the respondents had low BC and BSE knowledge. The basic BC knowledge was very low (2,47%); knowledge of BC symptoms, best time for BSE, and BSE associated activities was 9.96%, 4.58%, and 3.70% respectively. | There is need for a well-structured health education and screening programmes in Nigeria's educational institutions to enhance the prevention |
| 20 | 2024 | Udo, S. M; Ene, F. E; Ikechukwu, A. V; Gamde, S. M; Yetunde, E. A; Ogundipe, A. T; Oyedun, B. D; Nnenna, A. G; Bali, K. I; Biya, A; Inyang, A. E; Ogah, H. I. | Knowledge and associated risk factors of breast cancer among females in Karu local government area, Nasarawa State, Nigeria | Investigate knowledge and associated risk factors of breast cancer among females in Karu local government area, Nasarawa State, Nigeria | Nigeria | 300 female students | Cross sectional | There was a poor knowledge of breast cancer and associated risk factors among the study participants. | Study advocated for breast cancer awareness campaigns and accessible screening services. |

One was a multi-country study conducted in 24 low, middle income and emerging economy countries, including six (6) SSA countries (Cameroon, Ivory Coast, Madagascar, Mauritius, Namibia and Nigeria). The sample sizes in the studies included in the review ranged significantly, from as small as 89 (ID 18) to as large as 2,126 (ID 4). Sixteen (73%) of the study designs were mainly cross-sectional, followed by four (18%) descriptive one (1) pre-and post-interventional and one (1) longitudinal cohort study.

### Application of “awareness” and “knowledge” concepts

Assuming a subtle but important difference of concepts of “awareness” and “knowledge”, where “awareness” emphasizes being generally informed about breast cancer and its risk factors, and “knowledge” implies a more detailed and specific understanding of the breast cancer and its risk factors, we assessed the extent to which these two concepts were applied in the reviewed artcles. Forty percent (n=8) of the reviewed articles were of studies that included “awareness” only (IDs 2; 3; 4; 6; 7; 10; 12 and 16) in the title. An equal number (n=8) of articles also included “knowledge” only (IDs 5; 9; 11; 13; 14; 15; 17 and 20) in the titles. A fifth (n=4) of the articles included both “awareness” and “knowledge” (IDs 1; 8; 18 and 19) in the titles.

It was noted that regardless of the inclusion of the terms “awareness” or “knowledge” or both in the titles, authors frequently used “awareness” and “knowledge” interchangeably in most (n=17; 85.00%) reviewed articles. In one article, authors used the term “awareness” only in the main text (ID 4) and in one other article (ID 13), authors used the term “knowledge” only. In article ID 4, “level of awareness of breast cancer risk factors” was assessed with eight (8) questions among 2,126 female university students of six SSA countries (Cameroon, Ivory Coast, Madagascar, Mauritania, Namibia and Nigeria).

Authors for article ID 13 assessed “level of knowledge of breast cancer risk factors” with knowledge specific variables in a study conducted among 143 female students at Universtiy Uyo, Nigeria. One article (ID 19) of a study conducted in south western Nigeria, assessed the level of “awareness” using three closed ended structured questions and the level of “knowledge” using 14 close-ended structured questions. Nevertheless, the two terms were neither explicitly defined nor used interchangeably in article ID 19. These findings suggest a lack of distinction between the two concepts in most articles reviewed in this scoping review. Since the authors of the articlces included in this scoping review examined studies without a strict distinction between awareness and knowledge, we report our findings in the context of the combined levels as “awareness/knowledge” below.

### Awareness and knowledge levels of breast cancer, symptoms and signs

The first research question was on determining the nature and extent of research on on knowledge and awareness about breast cancer, its symtoms and signs. Seventy-five percent (n=15) of reviewed articles reported that breast cancer awareness/knowledge were high among participants, with rates varying significantly, from 58% to 100%. A limited number of articles reviewed (ID1, ID10, ID 12 and ID15) reported low levels of awareness/knowledge of breast cancer among females aged 10-24 years in SSA. Regarding signs/symptoms of breast cancer, levels of awareness/knowledge among the same age group in SSA were reported in 40% (n=8) of the articles included in the scoping review.

The review of twenty articles revealed a low proportion, with only three articles showing high levels of awareness about breast cancer signs/symptoms. The three articles reporting on high levels of awareness/knowledge of breast cancer signs/symptoms are ID3 at 78%, ID13 at 55% and ID20 at 74%. Five (5) articles reported levels below 50% (ID1, ID7, ID8, ID15 and ID19), and the rest did not report on awareness or knowledge of signs and symptoms of breast cancer among females aged 10-24 years in SSA. The most frequently and accurately reported signs and symptoms in the reviewed articles are pain in the breast region, changes in the breast’s morphology (shape and size), and nipple discharge.

### Awareness and knowledge levels of breast cancer risk factors

The second research question was on identifying research on knowledge and awareness about breast cancer risk factors among females aged 10-24 years in SSA. In our scoping review, most (n=14; 70%) of the articles reviewed showed a widespread lack of awareness/knowledge on risk factors associated with breast cancer, with a score of less than 50% (ID1, ID2, ID4, ID6, ID7, ID8, ID9, ID10, ID12, ID13, ID14, ID15, ID19 and ID20). Among these 14 articles, the most significant finding was revealed in one study conducted in northwest Ethiopia (ID 8) in which most female medical students were not aware of the complex risk factors associated with the breast cancer.

Additionally, other studies (ID10 and ID15) exposed low level of awareness/knowledge of breast cancer risk factors prior to an intervention (training, campaigns, symposium, reading handouts). Subsequent to the intervention, the studies exhibited significant positive changes in knowledge regarding the risk factors, encompassing most areas of understanding. However, in other six studies included in this scoping review, findings varied, ranging from “fair” (ID 18) to 100% (ID 3). Other descriptions used include “moderate” (ID 16), “intermediate” (ID 5), “satisfactory” (ID 11) and “most” (ID 17). The most cited risk factor in most of the articles included in this scoping review was “family history”.

### Awareness and knowledge of breast cancer screening methods

The third research question was on documenting evidence on levels of knowledge and awareness about breast cancer screening methods (BSE, CBE and mammography) among females aged 10-24 years in SSA. Almost all (95%) included articles reported on BSE awareness/knowledge levels (except articles ID4 and ID20), of which 30% of studies (ID6, ID7, ID10, ID12, ID14 and ID18) reported BSE awareness/knowledge levels below 50%. Moderate awareness/knowledge levels of BSE were reported in seven studies, of which three studies (ID15, ID16 and ID19) reported levels of 61%-70% and four studies (ID1, ID5, ID13 and ID17) reported levels between 71% and 80%. High awareness/knowledge in some SSA countries (ID2, ID3, ID11) indicated BSE awareness levels above 91%.

In the same set of 20 retrieved articles, 6 studies (30%) also investigated CBE, with their reported CBE awareness levels ranging from 7% (ID18) to 65% (ID3 and ID11). The remaining 14 articles discussed reported awareness levels of 17% (ID6), 36% (ID8) and 51.6% (ID16). Regarding mammography, awareness level was mentioned in 6 of the 20 studies reviewed, with participants level of awareness ranging from a low of 2.2% (ID18) to a high of 75.3% (ID11). Other studies cited levels of 5% (ID1), 34% (ID6), 65% (ID3) and 71.1% (ID16).

### Sources of information

The fourth research question focused on identifying the major sources of information through which females aged 10-24 years acquire breast health-related information in SSA. We found that 55% (n=11) of the articles reviewed reported different sources of information on various aspects of breast cancer. The most significant sources of information that facilitate breast cancer awareness/knowledge reported in the articles reviewed was media (social, electronic and print). Health workers ((ID14 and ID19), peer education and lectures (ID15 and ID6), parents (ID6 and ID10) and public campaigns (ID6) were noted as other sources of information. Additionally, most of these sources of information used in combination to create awareness/knowledge among AGYW in SSA during the reporting period.

### Strategies recommended in the reviewed articles

The fifth research question explored interventions recommended for improving knowledge and awareness of breast cancer among females aged 10-24 years in SSA. All the 20 reviewed articles emphasized the importance of increasing awareness/knowledge of breast cancer, symptoms, screening methods and risk factors at a young age. The reviewed studies highlight the need to educate women about the disease, including its risk factors, symptoms, and the importance of early detection. One other significant recommendation made in three articles reviewed (ID9, ID10 and ID18) was integration of breast health education into school curricula. The studies reviewed suggested reviewing existing materials and incorporating new content to teach students about breast health, risk factors, screening tools, and the importance of regular screenings.

Also, the positive impact of health education on awareness was demonstrated in two articles (ID10 and ID15) in Ngierian schools. Article ID10 showed a significant improvement on most aspects of knowledge regarding the breast cancer’s early symptoms and risk factors after the introduction of a six-months longitudinal intervention. In article ID10 used video-assisted face-to-face and printed handouts to create awareness and impart knowledge to adolescent female students. As a result of the intervention, they recommended the inclusion of breast cancer instructions into high school curricula. Similar results were also observed in arcticle ID15 with the use of a peer-education approach in a pre-post interventional study conducted among Nigerian female students at four secondary schools in Benin City. Article ID15 established that knowledge on breast cancer had improved from a score of 20.6% prior to peer-education to a high of 55.9% after peer-education (p < 0.0001), and recommended a low-cost strategy in low resource settings such as SSA countries.

## Discussion

The overlap in the definitions of knowledge and awareness necessitates precision for meaningful and useful differentiation of the two in public health research. Service providers can create awareness with the aim of improving the general knowledge of people about the risk factors of a public health condition. In this case, awareness is not the same as knowledge, but is simply creating a platform for people to understand facts on breast cancer, its risk factors, signs, symptoms and screening methods. Further, awareness often comes first, as individuals may have heard of breast cancer, while knowledge requires deeper learning and understanding of the details of the disease. While general awareness might encourage screening, lack of detailed knowledge may hinder informed decision-making about risk management and treatment options. Though the two terms can be used interchangeably in some cases, understanding the distinctions between knowledge and awareness is crucial for research contexts, especially when assessing public health interventions where precise measurement is needed. Therefore, this finding potentially leads to ambiguity in conveying meaning and has significant implications for breast cancer prevention and management.

Assessing general awareness and knowledge about breast cancer, its symptoms, and screening practices lays a strong foundation for conducting more in-depth research on specific risk factors. Our analysis reveals that the terms ‘awareness’ and ‘knowledge’ were frequently used interchangeably in the reviewed articles, hindering a deeper understanding of the concepts in the context of breast cancer, symptoms, associated risk factors and screening methods. The difference between breast cancer awareness and knowledge is fundamental for public health because both are distinct yet related, and they both contribute to early detection and better outcomes(World Health Organization, 2024)(World Health Organization, 2008).

The scoping review highlights a significant lack of research into awareness/knowledge of breast cancer, symptoms, risk factors, and screening in young women (10-24 years) across most of SSA, with studies predominantly limited to Nigeria. This research gap means a large portion of the region has not been studied for these critical breast health factors. This situation is exacerbated by by cultural factors, low health literacy, lack of comprehensive screening programs, insufficient health infrastructure, limited access to advanced therapies, and poor disease surveillance, all contributing to late diagnosis and high mortality rates in the region. general issues in the region like l, provider education gaps, and (Pace & Shulman, 2016). The limited research in the region hinders our ability to understand and address this significant health issue in this age group.

Our study indicates that in SSA, a significant portion of studies (75% of 20) found high breast cancer awareness among females aged 10-24 years. While this shows positive trends in breast cancer awareness among young women in SSA, this finding is based on limited geographic representation (15 out of 48 countries). The underlying message highlights a general trend of high awareness in some areas but points to an uneven distribution of research, with many countries in the region lacking sufficient studies to give a complete picture of breast cancer knowledge among young women. However, this high level of knowledge of breast reported among this age group is consistent with findings of similar studies conducted in other countries outside the SSA region, such as Gaza(Abo Al-Shiekh et al., 2021), Saudi Arabia(Alsareii et al., 2020), Egypt(Ahmed & Aljaber, 2018) and Nepal(Bishnu., 2023).

Our study results disclose a concerning lack of awareness and knowledge of breast cancer symptoms and risk factors among females aged 10-24 years in SSA. While there’s some general awareness and knowledge of breast cancer among young women aged 10-24 years, understanding of specific symptoms/signs and risk factors is limited in many parts of SSA. This scoping review highlights limited awareness and knowledge of breast cancer screening methods among young females (10-24 years) in SSA, with participants showing better knowledge of BSE than CBE or mammograms. Despite this, many women lack the practical skills for BSE, and overall knowledge about screening remains a significant challenge, compounded by limited access to services and cultural factors like stigma in the region. Given the limitations of mammography in low-resource settings, there is a growing emphasis on context-specific early detection strategies that are more feasible and culturally appropriate for the SSA.

This is consistent with findings from other parts of the world, outside SSA region. For instance, in a survey of 6380 Saudi Arabian female secondary school students, awareness of BSE was limited (39.6%), and the students’ understanding of breast cancer risk factors was also low(Milaat, 2000). This was observed in the Peoria, Illinois (USA) in a study involving ninth-grade female students from four public high schools(Ogletree et al., 2004). Another study conducted in Colombo, Sri Lanka, also revealed substantial knowledge gaps regarding breast cancer detection, services, and screening practices. Only 9.4% knew early detection methods, 5.5% knew where to seek help, 17.1% knew BSE procedures, and only 6.17% reported attempting it(Ranasinghe et al., 2013). Other studies with similar findings were conducted in Malaysia(Al-Naggar et al., 2011), Turkey(Karayurt et al., 2008) and northwestern India(Yadav & Jaroli, 2010).

This scoping review has identified studies that demonstrated significant improvements in public knowledge regarding breast cancer, symptoms and risk factors after evidence-based educational interventions in school settings. This improvement highlights the effectiveness of awareness campaigns and educational programs in enhancing public understanding of cancer signs and symptoms. Literature has shown that raising awareness and knowledge levels are the first steps in breast health care. Isara and Ojedokun (2011) argue that reduction in breast cancer deaths can only be facilitated through “health education” and “breast screening”(Isara & Ojedokun, 2011). They further contend that health education empowers women to be “breast aware”, teaching them the risk factors and early symptoms of breast cancers for better prognosis. In addition, it has been demonstrated that awareness creation and knowledge management encourage women to avoid risk factors and motivate prevention behavior(Bello, 2012).

Media can be a powerful tool for reaching adolescents with information on breast cancer due to its ability to disseminate information widely, facilitate peer-to-peer support, and offer a convenient way to access health resources. In this review, media (social, electronic and print) was cited as major source of information on breast cancer in this review. While social media can offer a platform for peer support, it’s important to remember that this support may not always be medically sound or appropriate. Social media can be a double-edged sword, with the potential for misinformation and lack of privacy. It is important to have a balanced approach that includes credible medical professionals, reputable health organizations, and peer support from trusted sources. This will provide a platform for females aged 10-24 years to develop a well-rounded understanding of breast cancer and make informed decisions about their health.

This age group is crucial for early detection and awareness initiatives, and educational programs are essential for promoting these practices. Furthermore, socioeconomic, cultural, psychological, misconceptions, stigma and false beliefs about breast cancer can create barriers to accurate knowledge and understanding, hindering this age group’s ability to make informed decisions about their well-being. Our review revealed a dearth of information on influence of these factors in knowledge of females aged 10-24 years, highlighting a critical gap in understanding the barriers this age group faces in accessing breast cancer information in SSA.

### Strenghts and limitations

This scoping review was conducted using a rigorous, standardized approach based on the Arksey and O’Malley framework for scoping reviews(Arksey & O’Malley, 2005), and provided an overview of the research landscape on breast cancer awareness and knowledge among females aged 10-24 years in SSA. Despite the highly rigorous methodology used in this study, inherent limitations that must be acknowledged to provide context, ensure transparency, and allow for proper interpretation of the research findings. For instance, the literature search was restricted to English language, and hence the possibility that we may have missed non-English publications that could have been crucial for drawing conclusions. However, emerging evidence suggests that this previously identified limitation is becoming less of a concern, as new findings indicate it does not have a substantial impact on the overall conclusions(Morrison et al., 2012).

### Implications for research and practice

While this review offers significant insights, it also identifies key areas for future investigation. Further research should explore the specific drivers of low breast cancer knowledge and awareness across diverse contexts within SSA. A standardized conceptualization of “knowledge” and “awareness” is also necessary to guide effective public health strategies. In light of the importance of early detection, our findings underscore the urgent need for targeted educational interventions in secondary and tertiary institutions to address the current knowledge gap.

### Conclusion

The study reveals limited research and low awareness/knowledge of breast cancer risk factors, symptoms/signs and screening practices among females aged 10-24 years in SSA, despite the rising burden of the disease in the region. These findings, while constrained by limited data, signal a critical need for targeted public health campaigns and educational programs to empower young women with knowledge about symptoms, risk factors, and screening to improve early diagnosis and outcomes.

## Data Availability

All relevant data are within the manuscript and its Supporting Information files.

## Contributions

All authors contributed to the development of the selection criteria, the quality assessment strategy, data extraction criteria, drafted the manuscript, read, provided feedback, and approved the final manuscript.

## Ethical Approval

This manuscript is a scoping review and is part of the outputs of the PhD thesis. The ethical approval for the PhD study was obtained from the Health Research Development Committee (HRDC) of the Ministry of Health (MOH), Botswana with ethical application reference number HPDME 13/18/1, as well as University of KwaZulu-Natal (UKZN) Biomedical Research Ethics Committee (BREC), South Africa, ethical application reference number BREC/00001596/2020. The approval letters are attached.

